# Development and Validation of a Machine Learning–Based Model for Predicting 6-Month Functional Outcomes in Patients with Intraventricular Hemorrhage Using the Brainstem Dorsal Line: A Multicenter Retrospective Cohort Study

**DOI:** 10.64898/2026.07.28.26359173

**Authors:** Chuhua Fu, Sai Du, Yuhang Zhao, Liansheng Mou, Shunan Hu, Juntao Hu, Yuan Yao, Xiong Wang, Yong Wu, Yi Huang, Songpu Li, Rong Hu, Hua Feng

**Author notes:** Correspondence to: Chuhua Fu, Department of Neurosurgery, Jingmen People’s Hospital, Jingchu University of Technology Affiliated People’s Hospital, Jingmen, China, E-mail address; Songpu Li, Intelligent Information Technology Research Center, Jingchu University of Technology, Jingmen 448000, China, E-mail address; Rong Hu, Department of Neurosurgery, Southwest Hospital, Army Medical University, Chongqing 400038, China, E-mail address; Hua Feng, Department of Neurosurgery, Southwest Hospital, Army Medical University, Chongqing 400038, China. Contributed equally and joint first authors (Chuhua Fu, Sai Du, Yuhang Zhao). Joint last authors (Chuhua Fu, Songpu Li, Rong Hu, Hua Feng).

## Abstract

**Objective:** Intraventricular haemorrhage (IVH) carries high mortality and morbidity; accurate early prediction of 6-month functional outcome is essential for guiding treatment decisions and optimising prognosis. We developed and externally validated a prognostic model that integrates a novel neuroimaging marker, the Brainstem Dorsal Line (BSDL), with the TabICLv2 algorithm to predict 6-month functional outcomes after IVH.

**Methods:** In this multicentre retrospective study, patients with IVH were enrolled from nine tertiary centres in China between Dec 1, 2020, and Dec 31, 2024; seven centres constituted the derivation cohort (8:2 training–validation split), and two centres formed the external validation cohort. Feature selection followed a three-step pipeline comprising univariable testing, Spearman correlation analysis, and LASSO regression; eight machine-learning models were trained with five-fold cross-validated hyperparameter tuning. Performance was assessed by the area under the receiver operating characteristic curve (AUC), the area under the precision–recall curve (AUPRC), calibration curves, and decision curve analysis (DCA). Interpretability was evaluated using SHapley Additive exPlanations (SHAP).

**Results:** In total, 728 eligible patients were enrolled — 610 in the derivation cohort (unfavourable outcome, 27·9%) and 118 in the external validation cohort (26·3%). Feature selection identified nine optimal predictors, of which BSDL grade 2 ranked highest in feature importance. TabICLv2 showed the best performance in external validation (AUC 0·9014 [95% CI 0·825–0·985], AUPRC 0·8192 [0·688–0·916], F1 score 0·7742 [0·651–0·877]); calibration was excellent (Hosmer– Lemeshow test, P>0·05) and DCA showed clinical net benefit across threshold probabilities of 0·05–0·97. SHAP confirmed BSDL grade 2 as the strongest predictor of unfavourable outcome.

**Conclusions:** Integrating BSDL with TabICLv2 yielded a robust, interpretable prognostic model for 6-month outcomes after IVH, providing a practical tool for early risk stratification and individualised treatment planning.

## 1. Introduction

Spontaneous intracerebral haemorrhage (sICH) is among the stroke subtypes with the highest mortality and disability, accounting for 10–20% of all strokes and ranking as the second leading cause of stroke-related death globally^1–6^. Intraventricular haemorrhage (IVH), a severe complication affecting 20–60% of sICH patients, carries a 30-day mortality of 40–60%, and only 25% of survivors achieving functional independence at 6 months^2,6–8^. Since neurological recovery occurs predominantly within the first 3–6 months, accurate early prognostic assessment is critical for risk stratification, individualised treatment, and rational resource allocation^9^.

Conventional prognostic indicators — including Glasgow Coma Scale (GCS), haematoma volume, hydrocephalus, and original/modified Graeb scores (oGS/mGS) — derive largely from univariate analyses and inadequately capture the multifactorial pathophysiology of IVH, thus limiting predictive performance^10–11^. Advances in radiomics and artificial intelligence have spurred the use of machine learning (ML) to integrate multidimensional clinical and imaging data, yielding more accurate prognostic models^4,11–12^. ML has been widely applied to outcome prediction in cerebrovascular disease, including sICH, subarachnoid haemorrhage, and ischaemic stroke ^3,9,13–18^. Nevertheless, significant gaps remain. No prognostic model has been developed specifically for IVH, with most studies merely classifying its presence as a binary variable. Although fourth-ventricle haematoma volume is an independent prognostic risk factor^8,10,19–21^, no standardised quantitative or semi-quantitative framework exists for assessing its compressive effect on brainstem structures. Moreover, existing models predominantly rely on conventional algorithms (e.g., logistic regression, random forest) that fail to adequately capture complex variable interactions in tabular clinical data; many also lack rigorous external validation and interpretability, hindering clinical translation^3,9,13,21^.

To address these limitations, we propose the Brainstem Dorsal Line (BSDL), a novel imaging grading concept that quantifies brainstem compression caused by fourth ventricular haematoma. Building on our previous finding that ventricular haematoma burden is a key determinant of IVH prognosis^22^, we applied the Tabular In-Context Learning version 2 (TabICLv2) algorithm, a deep learning model optimised for tabular data^23^, to develop and externally validate the first interpretable model that predicts 6-month functional outcomes in a multicentre IVH cohort. By integrating BSDL grading with the TabICLv2, we aim to develop an accurate, interpretable prognostic tool that can refine risk stratification and inform individualised management for patients with IVH.

## 2. Methods

### 2.1 Study design and patients

In this multicentre retrospective cohort study, we consecutively enrolled patients with IVH admitted to nine tertiary medical centres in China between Dec 1, 2020, and Dec 31, 2024. Patients from seven centres (Dec 1, 2020–Dec 31, 2023)formed the derivation cohort and were randomly split into training and internal validation sets in an 8:2 ratio. The remaining two centres (Jan 1, 2024–Dec 31, 2024) comprised the independent external validation cohort. Sample size met the recommended events-per-variable (EPV) ratio of ≥10 for clinical prediction model studies^2,12,17^.

Inclusion criteria were: (1) age 18–80 years; (2) intensive care unit admission; (3) primary IVH or secondary IVH due to sICH (parenchymal haematoma volume ≤20 ml); (4) pre-onset modified Rankin Scale (mRS) score of 0 or 1. Exclusion criteria were: (1) secondary haemorrhage from intracranial aneurysm, arteriovenous malformation, tumour-related stroke or other aetiologies; (2) coexisting choroid plexus vascular malformation or moyamoya disease; (3) systemic coagulopathy; (4) platelet count <100 × 10⁹/L or international normalised ratio (INR) >1·4; (5) pregnancy; (6) concomitant brainstem haemorrhage; (7) previous craniotomy; (8) active bleeding (retroperitoneal, gastrointestinal, urinary, respiratory, or other sites) before admission or recent surgery; or (9) incomplete medical records, missing imaging data, or loss to follow-up. Diagnosis and treatment adhered to the 2022 AHA/ASA Guideline for the Management of Patients With Spontaneous Intracerebral Haemorrhage and the 2019 Chinese Guidelines for the Diagnosis and Treatment of Intracerebral Haemorrhage^1,24^.

The study was approved by the ethics committees of the lead centres (KY2023149, 2022KY065) and conducted in accordance with the Declaration of Helsinki. Given the retrospective design, the requirement for informed consent was waived. The study was registered on medicalresearch.org.cn (MR-50-23-048489) and reported according to the TRIPOD+AI statement^25–26^ . The detailed study workflow is presented in Figure 1.

**Figure 1:**
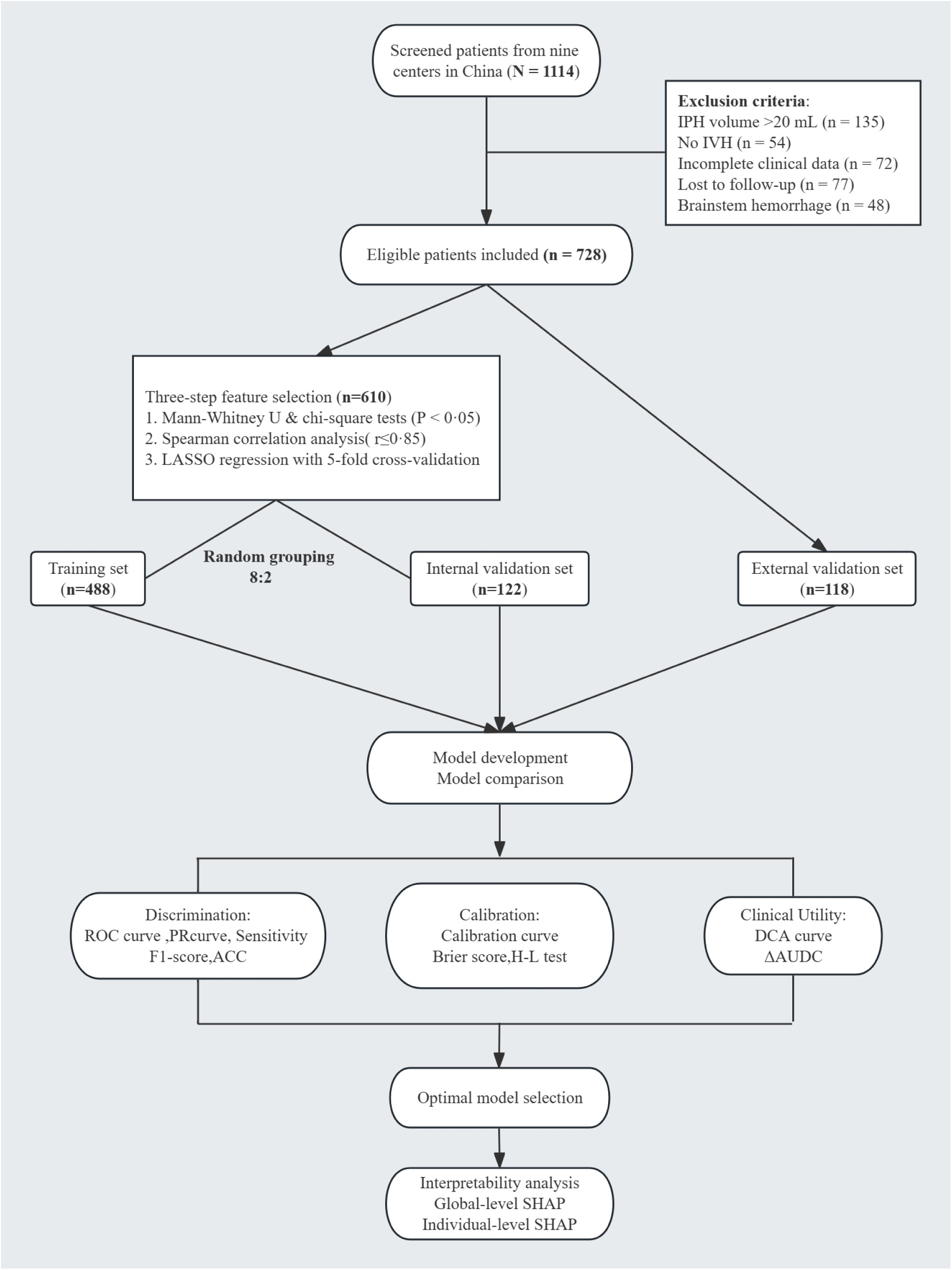
Flowchart of this study. IPH=intraparenchymal haemorrhage. IVH=intraventricular haemorrhage. LASSO=least absolute shrinkage and selection operator. ROC=receiver operating characteristic. PR=precision recall. ACC= accuracy. H-L=Hosmer-Lemeshow. DCA=decision curve analysis. ΔAUDC=incremental area under the decision curve. SHAP=SHapley Additive exPlanations.

### 2.2 Definition and measurement of the key indicator: BSDL

The BSDL was measured on sagittal multiplanar reconstruction of the admission cranial CT scans. On the standard midsagittal plane, we traced the dorsal brainstem contour; the ventral curvature and maximal shift distance (SD) were recorded. SD was defined as the perpendicular distance from the point of greatest brainstem compression to the hypothetical straight reference line representing the uncompressed state (Figure 2K). Based on morphological features and SD, BSDL was classified into three grades:

**Figure 2:**
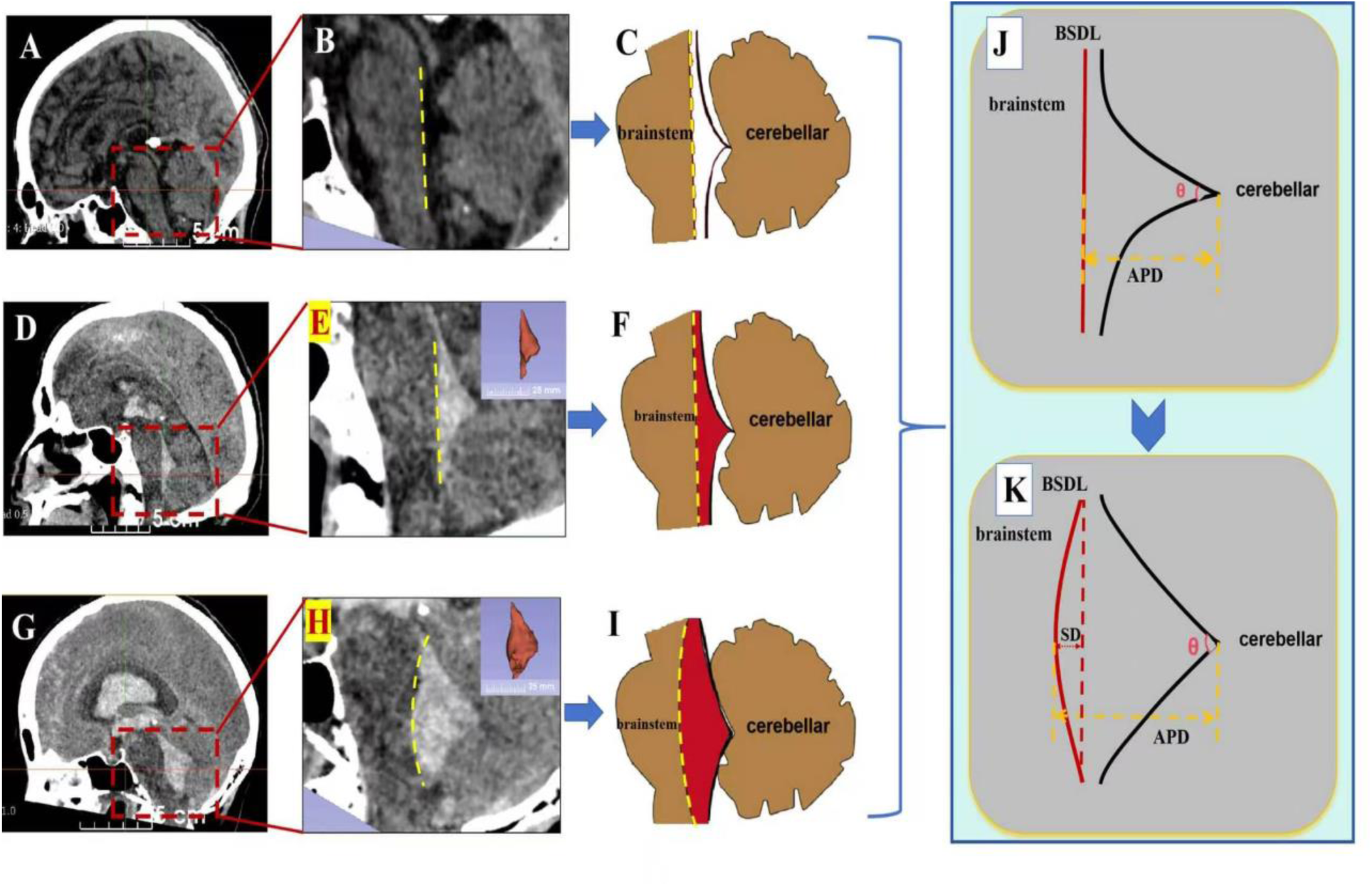
Morphological classification of fourth ventricular haemorrhage. (A–C) No haematoma. (D–F) Haemorrhage without ventricular expansion. (G–I) Haemorrhage with ventricular expansion. (J–K) Schematic diagrams of BSDL. All images were anonymized and no identifiable patient information is shown. BSDL=brainstem dorsal line. APD=anteroposterior diameter of the fourth ventricle. SD=shift distance.

BSDL grade 0: no or minimal haemorrhage in the fourth ventricle; the BSDL was straight and brainstem morphology intact (Figure 2A–C).

BSDL grade 1: haemorrhage was present in the fourth ventricle without obvious brainstem deformation; the BSDL was straight or showed only slight arcuate displacement, with SD ≤2 mm (Figure 2D–F).

BSDL grade 2 : a large fourth-ventricle haematoma caused ventral displacement of the brainstem; the BSDL showed marked arcuate displacement, with SD >2 mm (Figure 2G–I).

Two neuroradiologists, blinded to outcomes, independently assessed all scans; disagreements were resolved by a third senior neuroradiologist.

### 2.3 Data collection and outcome definition

At each centre, two trained neurosurgeons (≥5 years’ experience) independently extracted data; discrepancies were adjudicated by a senior neurosurgeon (≥10 years’ experience). All data handling complied with applicable privacy regulations. Collected variables comprised demographics, medical history, clinical and imaging characteristics on admission, and treatment modalities (Table 1). Haematoma volume was measured using the slice-by-slice tracing function of 3D Slicer (version 4.10.2; http://www.slicer.org)^8–9^.

**Table 1.** Data Fields Extracted from Electronic Medical Record.

| Category | Variables |
| --- | --- |
| Demographics | Age, sex |
| Past medical/medication history | Hypertension, DM, hyperlipidaemia , PH-APT , PH-ACT |
| Clinical characteristics on arrival | SBP, GCS , mGS , oGS |
| Radiological features at admission | Location of IPH, IPH volume, lateral IVH volume, third vent. IVH volume, fourth vent. IVH volume, BSDL, AH, ambient cistern, prepontine cistern |
| Treatment modalities | Conservative treatment, EVD placement,<br>Supratentorial haematoma evacuation + EVD<br>Infratentorial haematoma evacuation + EVD |
| Outcomes | Favourable outcome (mRS 0–3)<br>Unfavourable outcome (mRS 4–6) |

AH=acute hydrocephalus. BSDL=brainstem dorsal line. DM=diabetes mellitus. EVD=external ventricular drainage. GCS=Glasgow Coma Scale. IPH=intraparenchymal haemorrhage. IVH=intraventricular haemorrhage. mGS=modified Graeb score. mRS=modified Rankin Scale. oGS=original Graeb score. PH-ACT=prehospital anticoagulation therapy. PH-APT=prehospital antiplatelet therapy. SBP=systolic blood pressure. SHE=supratentorial haematoma evacuation. IHE=infratentorial haematoma evacuation. vent.=ventricular.

The primary outcome was functional functional status at 6 months after onset, assessed using the mRS via standardised telephone or face-to-face follow-up by assessors blinded to baseline data^27^. Consistent with prior studies, an mRS score of 0–3 was defined as a favourable outcome and 4–6 as an unfavourable outcome^16–17,20,26–28^.

### 2.4 Data preprocessing and feature selection

Raw CT images were standardized using 3D Slicer (version 4.10.2) for window width/level adjustment and resampling to isotropic voxels. Preprocessing included missing-value imputation (multiple imputation) and outlier correction; dichotomisation of age (<65 vs ≥65 years), SBP (<175 vs ≥175 mmHg), and GCS (3–8 vs 9–15); binarisation of haematoma volumes at Youden-index-optimised thresholds; and Z-score standardisation of continuous and ordinal encoding of categorical variables.

Feature selection proceeded in three steps: univariate filtering (Mann–Whitney U test or χ² test, P<0·05 retained); exclusion of collinear variables (Spearman correlation analysis, r>0·85); and LASSO regression with five-fold cross-validated, retaining non-zero-coefficient variables.

### 2.5 Model development and hyperparameter optimization

Using the optimal feature subset, we developed and systematically compared eight ML models: logistic regression (LR), random forest (RF), support vector machine with kernel (SVM Kernel), extreme gradient boosting (XGBoost), light gradient boosting machine (LightGBM), neural network (NN), tabular prior-data fitted network (TabPFN), and TabICLv2. These span classical linear, tree-ensemble, kernel-based, neural network, and tabular-specific deep-learning approaches. All models were trained with five-fold stratified cross-validation to improve performance and mitigate overfitting. To mitigate potential bias from class imbalance, class weights were set inversely proportional to class frequencies for models supporting this feature. Hyperparameters were optimised using grid search with manual fine-tuning.

### 2.6 Model evaluation and interpretability analysis

Model performance was evaluated across three dimensions: discrimination, calibration, and clinical utility.

#### Discrimination

Primary metrics were the area under the receiver operating characteristic curve (AUC) and the area under the precision–recall curve (AUPRC); secondary metrics included accuracy, sensitivity, and F1 score.

#### Calibration

Agreement between predicted probabilities and observed event rates was assessed using calibration curves, the Brier score, and the Hosmer–Lemeshow (H–L) test. Lower Brier score indicated better calibration, and a Hosmer–Lemeshow P>0·05 indicated no significant miscalibration.

#### Clinical utility

Decision curve analysis (DCA) was used to quantify clinical net benefit across a range of threshold probabilities, and the incremental area under the decision curve (ΔAUDC) was computed to quantify the clinical gain of each model relative to alternative strategies.

#### Interpretability analysis

To address the inherent “black-box” nature of ML, the best-performing model was interpreted using SHapley Additive exPlanations (SHAP). SHAP values were computed for each feature, and feature-importance bar charts, dependence plots, and individual waterfall plots were generated to elucidate model decision logic at both global and individual levels.

### 2.7 Statistical analysis

Statistical analyses were performed using SPSS version 27·0 and Python version 3·9. Continuous variables were first assessed for normality. Normally distributed variables were expressed as mean (SD) and compared using the independent-samples t test. Non-normally distributed variables were expressed as median (IQR) and compared using the Mann–Whitney U test. Categorical variables were expressed as n (%) and compared using the χ² test or Fisher’s exact test. All tests were two-sided, and P<0·05 was considered statistically significant.

## 3. Results

### 3.1 Patients’ characteristics

A total of 728 patients with IVH were enrolled: 488 in training, 122 in internal validation, and 118 in external validation cohorts. Unfavourable 6-month outcomes occurred in 27·9%, 27·9%, and 26·3% of patients, respectively (p>0·05).

Baseline characteristics were generally balanced across cohorts (Table S1), although the external validation cohort showed expected heterogeneity in admission SBP (P=0·001), mGS (P=0·001), lateral IVH volume (P=0·046), hyperlipidaemia (P=0·03), and location of IPH (P=0·01).

Within the derivation cohort, baseline characteristics by outcome are shown in Table 2 (favourable, n=440; unfavourable, n=170). The two groups did not differ significantly in age, sex, or medical history. However, patients with unfavourable outcomes had lower GCS, higher mGS and oGS, and larger haematoma volumes (all P<0·001). Notably, BSDL grade 2 was much more common in the unfavourable group (47·1% vs 4·5%; P<0·001).

**Table 2.**
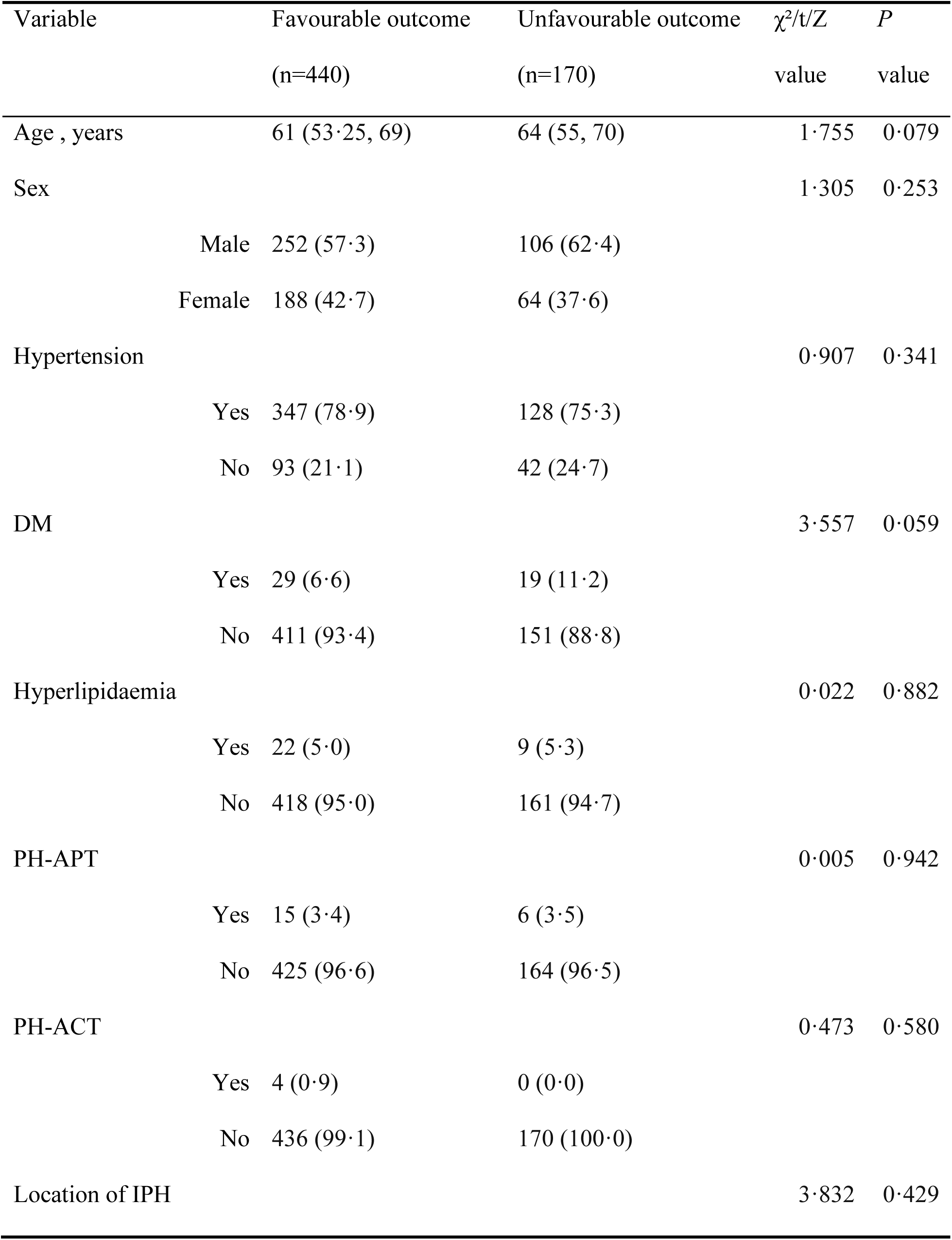
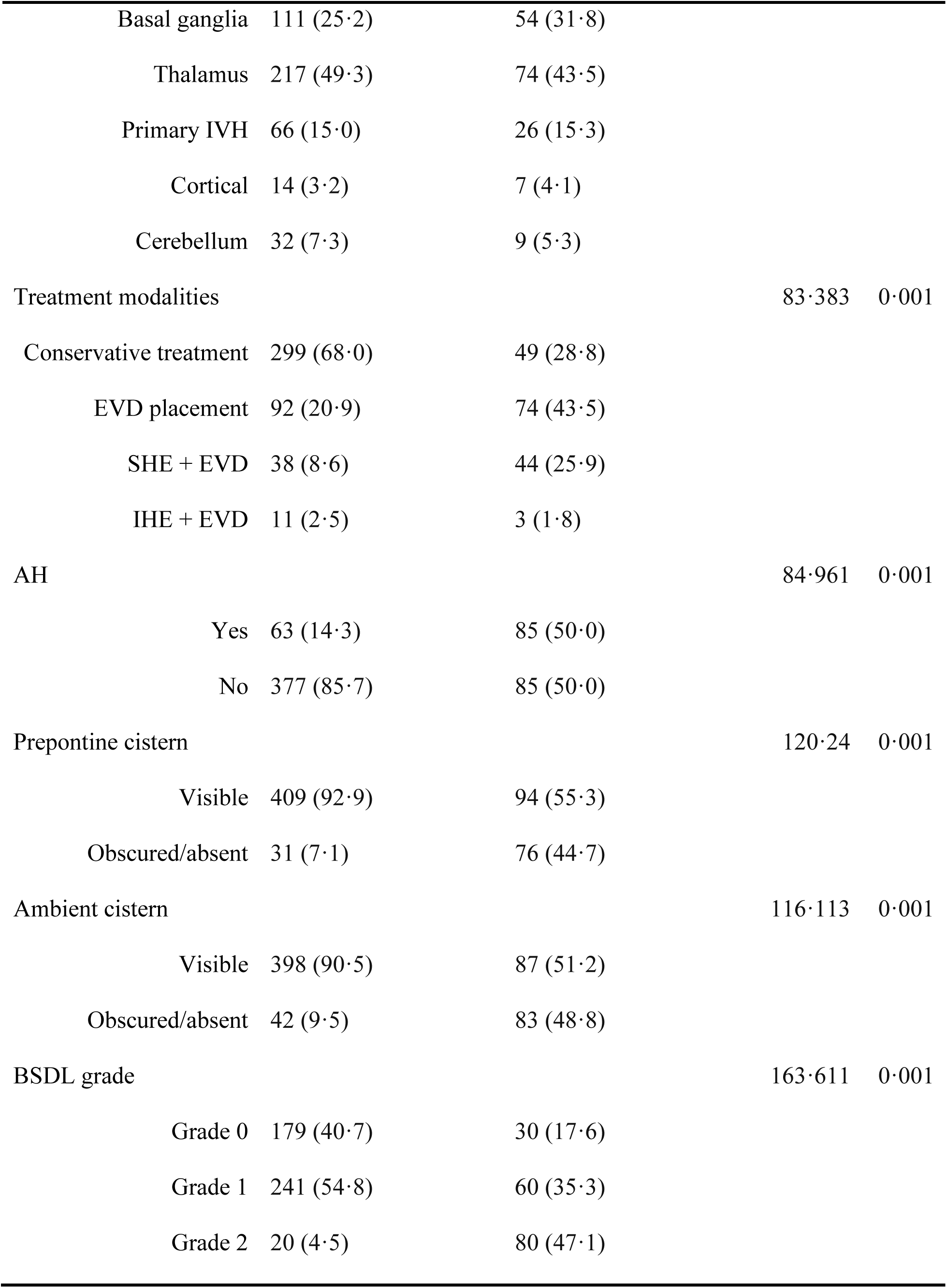
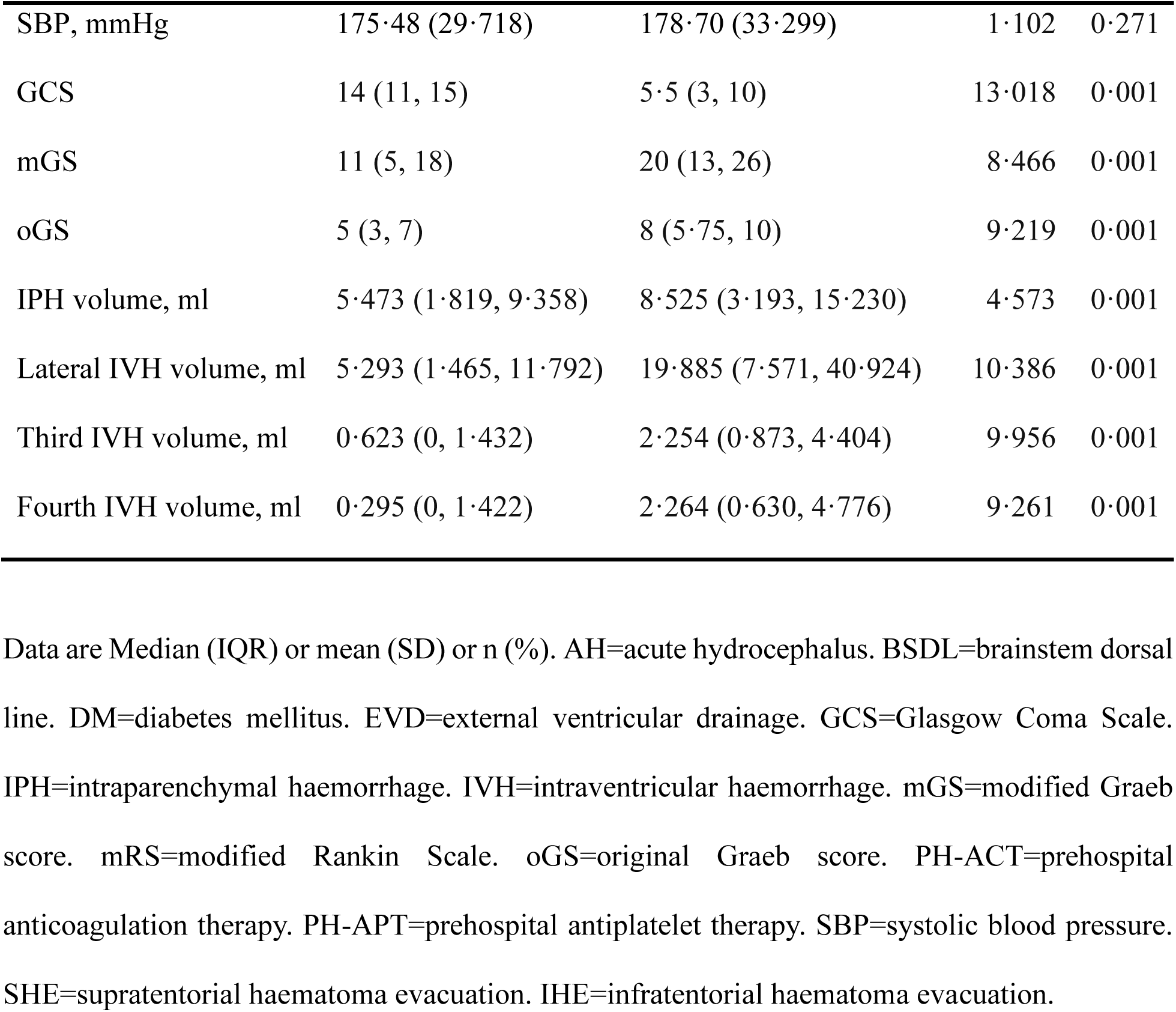
Baseline characteristics by 6-month functional outcome in the derivation cohort.

| Variable | Favourable outcome<br>(n=440) | Unfavourable outcome<br>(n=170) | $\chi^2/t/Z$<br>value | *P*<br>value |
| --- | --- | --- | --- | --- |
| Age , years | 61 (53.25, 69) | 64 (55, 70) | 1.755 | 0.079 |
| Sex |  |  | 1.305 | 0.253 |
| Male | 252 (57.3) | 106 (62.4) |  |  |
| Female | 188 (42.7) | 64 (37.6) |  |  |
| Hypertension |  |  | 0.907 | 0.341 |
| Yes | 347 (78.9) | 128 (75.3) |  |  |
| No | 93 (21.1) | 42 (24.7) |  |  |
| DM |  |  | 3.557 | 0.059 |
| Yes | 29 (6.6) | 19 (11.2) |  |  |
| No | 411 (93.4) | 151 (88.8) |  |  |
| Hyperlipidaemia |  |  | 0.022 | 0.882 |
| Yes | 22 (5.0) | 9 (5.3) |  |  |
| No | 418 (95.0) | 161 (94.7) |  |  |
| PH-APT |  |  | 0.005 | 0.942 |
| Yes | 15 (3.4) | 6 (3.5) |  |  |
| No | 425 (96.6) | 164 (96.5) |  |  |
| PH-ACT |  |  | 0.473 | 0.580 |
| Yes | 4 (0.9) | 0 (0.0) |  |  |
| No | 436 (99.1) | 170 (100.0) |  |  |
| Location of IPH |  |  | 3.832 | 0.429 |
| Basal ganglia | 111 (25·2) | 54 (31·8) |  |  |
| Thalamus | 217 (49·3) | 74 (43·5) |  |  |
| Primary IVH | 66 (15·0) | 26 (15·3) |  |  |
| Cortical | 14 (3·2) | 7 (4·1) |  |  |
| Cerebellum | 32 (7·3) | 9 (5·3) |  |  |
| Treatment modalities |  |  | 83·383 | 0·001 |
| Conservative treatment | 299 (68·0) | 49 (28·8) |  |  |
| EVD placement | 92 (20·9) | 74 (43·5) |  |  |
| SHE + EVD | 38 (8·6) | 44 (25·9) |  |  |
| IHE + EVD | 11 (2·5) | 3 (1·8) |  |  |
| AH |  |  | 84·961 | 0·001 |
| Yes | 63 (14·3) | 85 (50·0) |  |  |
| No | 377 (85·7) | 85 (50·0) |  |  |
| Prepontine cistern |  |  | 120·24 | 0·001 |
| Visible | 409 (92·9) | 94 (55·3) |  |  |
| Obscured/absent | 31 (7·1) | 76 (44·7) |  |  |
| Ambient cistern |  |  | 116·113 | 0·001 |
| Visible | 398 (90·5) | 87 (51·2) |  |  |
| Obscured/absent | 42 (9·5) | 83 (48·8) |  |  |
| BSDL grade |  |  | 163·611 | 0·001 |
| Grade 0 | 179 (40·7) | 30 (17·6) |  |  |
| Grade 1 | 241 (54·8) | 60 (35·3) |  |  |
| Grade 2 | 20 (4·5) | 80 (47·1) |  |  |
| SBP, mmHg | 175·48 (29·718) | 178·70 (33·299) | 1·102 | 0·271 |
| GCS | 14 (11, 15) | 5·5 (3, 10) | 13·018 | 0·001 |
| mGS | 11 (5, 18) | 20 (13, 26) | 8·466 | 0·001 |
| oGS | 5 (3, 7) | 8 (5·75, 10) | 9·219 | 0·001 |
| IPH volume, ml | 5·473 (1·819, 9·358) | 8·525 (3·193, 15·230) | 4·573 | 0·001 |
| Lateral IVH volume, ml | 5·293 (1·465, 11·792) | 19·885 (7·571, 40·924) | 10·386 | 0·001 |
| Third IVH volume, ml | 0·623 (0, 1·432) | 2·254 (0·873, 4·404) | 9·956 | 0·001 |
| Fourth IVH volume, ml | 0·295 (0, 1·422) | 2·264 (0·630, 4·776) | 9·261 | 0·001 |

Data are Median (IQR) or mean (SD) or n (%). AH=acute hydrocephalus. BSDL=brainstem dorsal line. DM=diabetes mellitus. EVD=external ventricular drainage. GCS=Glasgow Coma Scale. IPH=intraparenchymal haemorrhage. IVH=intraventricular haemorrhage. mGS=modified Graeb score. mRS=modified Rankin Scale. oGS=original Graeb score. PH-ACT=prehospital anticoagulation therapy. PH-APT=prehospital antiplatelet therapy. SBP=systolic blood pressure. SHE=supratentorial haematoma evacuation. IHE=infratentorial haematoma evacuation.

### 3.2 Feature selection results

After the three-step feature selection, nine variables were retained: BSDL grade, GCS, IPH volume, lateral IVH volume, third-ventricle IVH volume, oGS, AH, mGS, and fourth-ventricle IVH volume.

### 3.3 Evaluation and comparison of model performance

Using this feature subset, we trained eight ML models and evaluated them across discrimination, calibration, and clinical utility. Performance metrics across the training, internal validation, and external validation sets are shown in Table S2.

#### 3.3.1 Discrimination

Discrimination was assessed using receiver operating characteristic (ROC) curves, precision– recall (PR) curves, and their corresponding areas under the curves (Figure 3). In the training set, TabPFN (AUC 0·8203 [95% CI 0·770–0·869]) and TabICLv2 (AUC 0·8181 [0·768–0·868]) exhibited superior discriminative performance, with higher AUPRCs than conventional models (Figure 3a,b). In the internal validation set, TabICLv2 maintained high performance (AUC 0·8900 [0·809–0·952], AUPRC 0·8155 [0·672–0·912]; Figure 3c,d). In the external validation set, TabICLv2 demonstrated robust generalization (AUC 0·9014 [0·825–0·958], AUPRC 0·8193 [0·688–0·916]) with balanced precision-recall performance, whereas the NN showed high AUC (0·9103 [0·855–0·954]) but markedly lower AUPRC (0·7985 [0·672–0·905]) and sensitivity(0·4839 [0·310–0·667]; Figure 3e,f).

**Figure 3:**
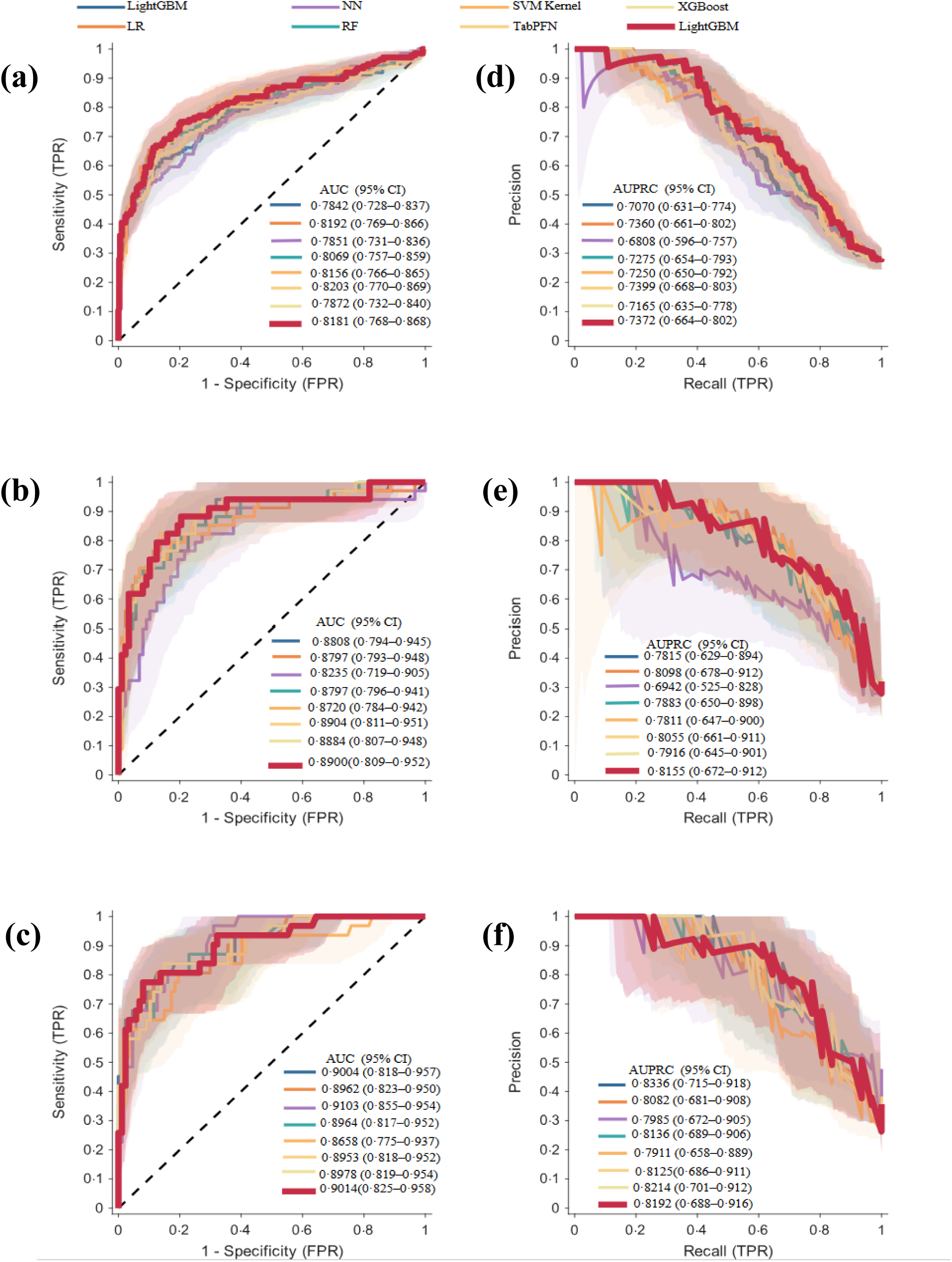
Discriminative performance of eight ML models across the training, validation, and external test sets. ROC curves are shown for the training set (a), validation set (b), and external test set (c). PR curves are shown for the corresponding datasets in panels (d), (e), and (f). The diagonal dashed line in (a–c) represents the reference line of no discrimination (AUC = 0·5). Shaded areas represent the 95% confidence intervals for each model. AUC values for ROC curves and average precision (AUPRC) values for PR curves are reported in the legends. AUC=area under the curve. AP=average precision. ML=machine learning. PR=precision-recall. ROC= receiver operating characteristic. FPR=false positive rate. TPR=true positive rate.

#### 3.3.2 Calibration

TabICLv2 demonstrated excellent calibration across all cohorts, with Brier scores of 0·1301 (95% CI 0·109–0·151), 0·1074 (0·074–0·146), and 0·0986 (0·067–0·139) in the training, internal validation, and external validation sets, respectively; calibration curves closely tracked the ideal diagonal (Figure S1). The Hosmer–Lemeshow test indicated no significant miscalibration in either the internal (P=0·587) or external (P=0·377) validation set.

#### 3.3.3 Clinical utility

Clinical utility was assessed by DCA, with net benefit and the incremental area under the decision curve (ΔAUDC) as primary metrics (Figure 4). TabICLv2 consistently yielded higher net benefit than all comparator models and both reference strategies (“treat-all” and “treat-none”) across all cohorts. Compared with the next-best model, TabICLv2 achieved ΔAUDC values of 0·0718, 0·0883, and 0·0908 in the training, internal validation, and external validation sets, respectively. These gains increased with cohort heterogeneity, suggesting robust generalisation without overfitting.

**Figure 4:**
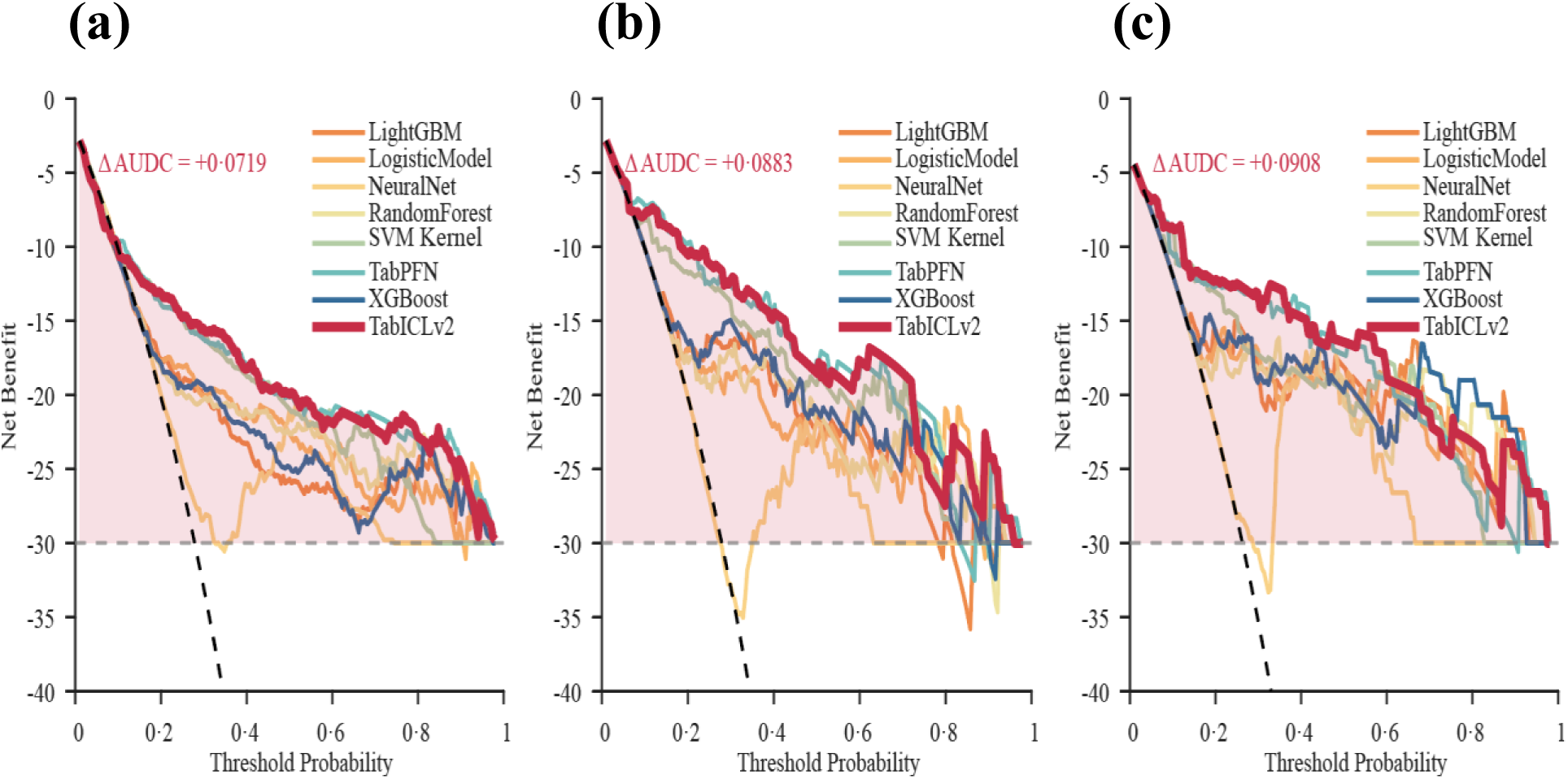
DCA comparing models in the training set (a), internal validation set (b), and external validation set (c). The x-axis indicates the threshold probability for clinical intervention, and the y-axis shows net benefit. The grey dashed line represents the treat-all strategy, and the black dashed line represents the treat-none strategy. The ΔAUDC for TabICLv2 relative to the next-best model is shown in the top left corner of each panel. DCA=decision curve analysis. ΔAUDC=incremental area under the decision curve.

#### 3.3.4 Classification Performance and Optimal Model Selection

Confusion matrices at optimal thresholds in the external validation set are shown in Figure S2.

TabICLv2 achieved the best balance for clinical decision-making: sensitivity 77·4%, specificity 92·0%, NPV 92·0%, and PPV 77·4%. By contrast, TabPFN achieved higher sensitivity (80·7%) but lower PPV (64·1%), whereas the NN achieved higher specificity (96·6%) at the expense of sensitivity (48·4%).

Based on combined evaluation of discrimination, calibration, and clinical utility, TabICLv2 was identified as the optimal model, demonstrating robust performance in the external validation set: AUC 0·9014 (95% CI 0·825–0·958), AUPRC 0·8192 (0·688–0·916), Brier score 0·0986 (0·067–0·139), sensitivity 0·7742 (0·618–0·909), accuracy 0·8814 (0·822–0·932), and F1 score 0·7742 (0·651–0·877). Performance degradation across cohorts was minimal, indicating the strongest cross-cohort consistency and generalisability among all models evaluated.

### 3.4 Interpretability Analysis

To elucidate the decision logic of TabICLv2, SHAP analysis was performed at global and individual levels in the external validation set. After ordinal encoding, GCS was represented as two binary features (GCS1: 9–15; GCS2: 3–8) and BSDL grade as three binary features (grades 0, 1, and 2), yielding 12 features for analysis (Figure 5).

**Figure 5.**
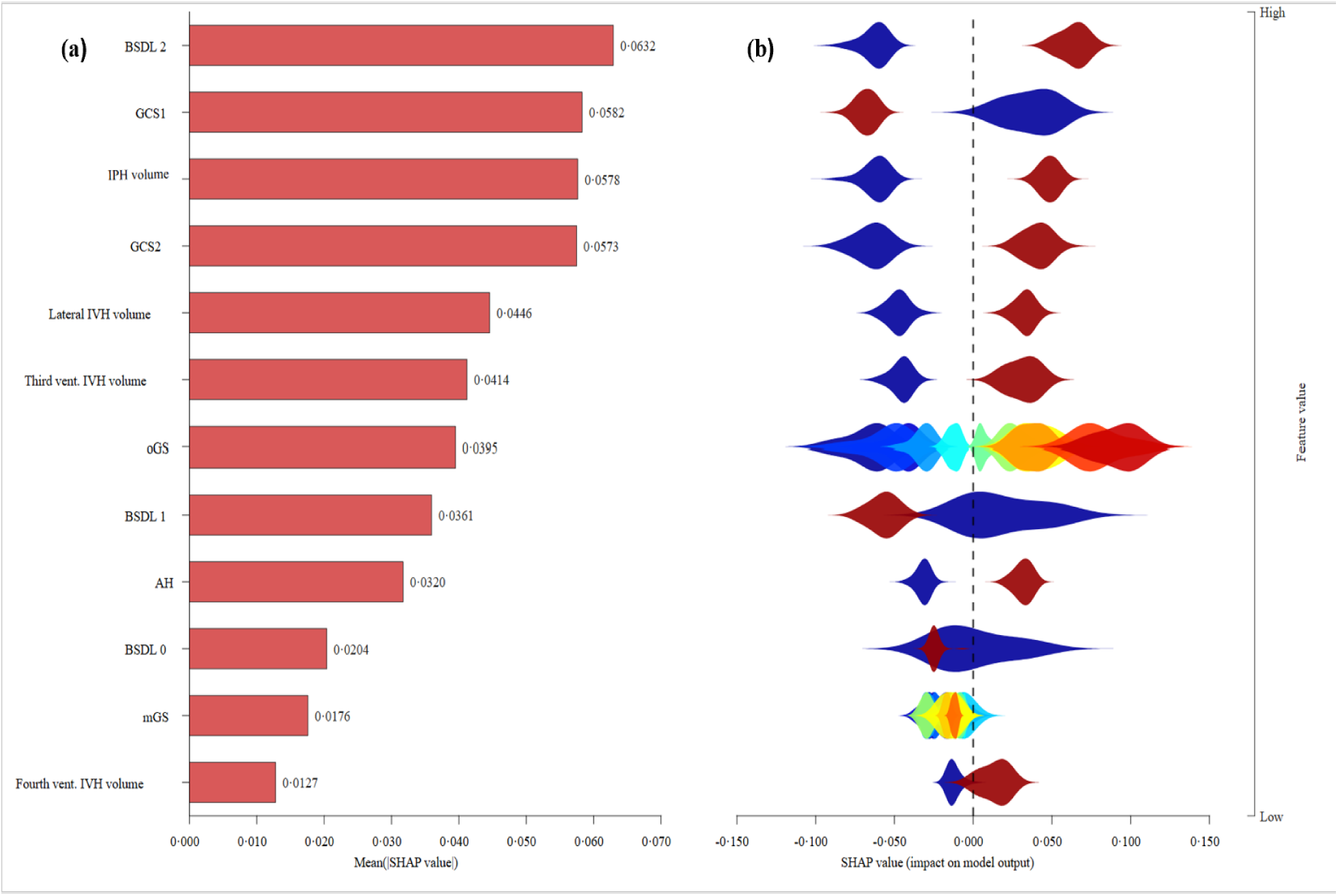
Global-level SHAP interpretability analysis of the TabICLv2 model in the external validation cohort. (a) Feature importance bar plot, ranked in descending order by mean absolute SHAP value. (b) SHAP violin plot: SHAP values on the x-axis (positive values indicate increased risk of poor outcome; negative values indicate decreased risk); colour gradient indicates feature values (blue=low, red=high); violin width corresponds to sample density at each SHAP value. AH=acute hydrocephalus. BSDL=brainstem dorsal line. GCS=Glasgow Coma Scale (GCS1, 9– 15; GCS2, 3–8). IPH=intraparenchymal haemorrhage. IVH=intraventricular haemorrhage. mGS=modified Graeb score. oGS=original Graeb score. vent.=ventricular.

#### 3.4.1 Global-level SHAP Interpretability Analysis

Global feature importance (Figure 5a), ranked by mean absolute SHAP value, identified BSDL grade 2 as the leading contributor (0·063), followed by GCS1 (0·058), IPH volume (0·058), and GCS2 (0·057). Secondary contributors included lateral IVH volume, third-ventricle IVH volume, oGS, BSDL grade 1, and AH, while BSDL grade 0, mGS, and fourth-ventricle IVH volume contributed minimally. This hierarchy indicates that brainstem injury, level of consciousness, and haematoma burden are the primary drivers of unfavourable outcome prediction.

The SHAP violin plot (Figure 5b) showed that BSDL grade 2, GCS2, greater haematoma burden, and higher oGS values carried positive contributions (increased predicted risk of unfavourable outcome), whereas BSDL grade 0 and GCS1 carried protective negative contributions. BSDL grade 2 exhibited the widest SHAP distribution (−0·100 to +0·150), with high-value samples potentially increasing predicted risk by up to 15%. Effect magnitude decreased stepwise from grade 2 to grade 0, confirming a dose–response relationship.

#### 3.4.2 Local-level SHAP Interpretability Analysis

Two representative cases from the external validation set were examined using SHAP waterfall plots to illustrate individual-level decision logic (Figure 6).

**Figure 6:**
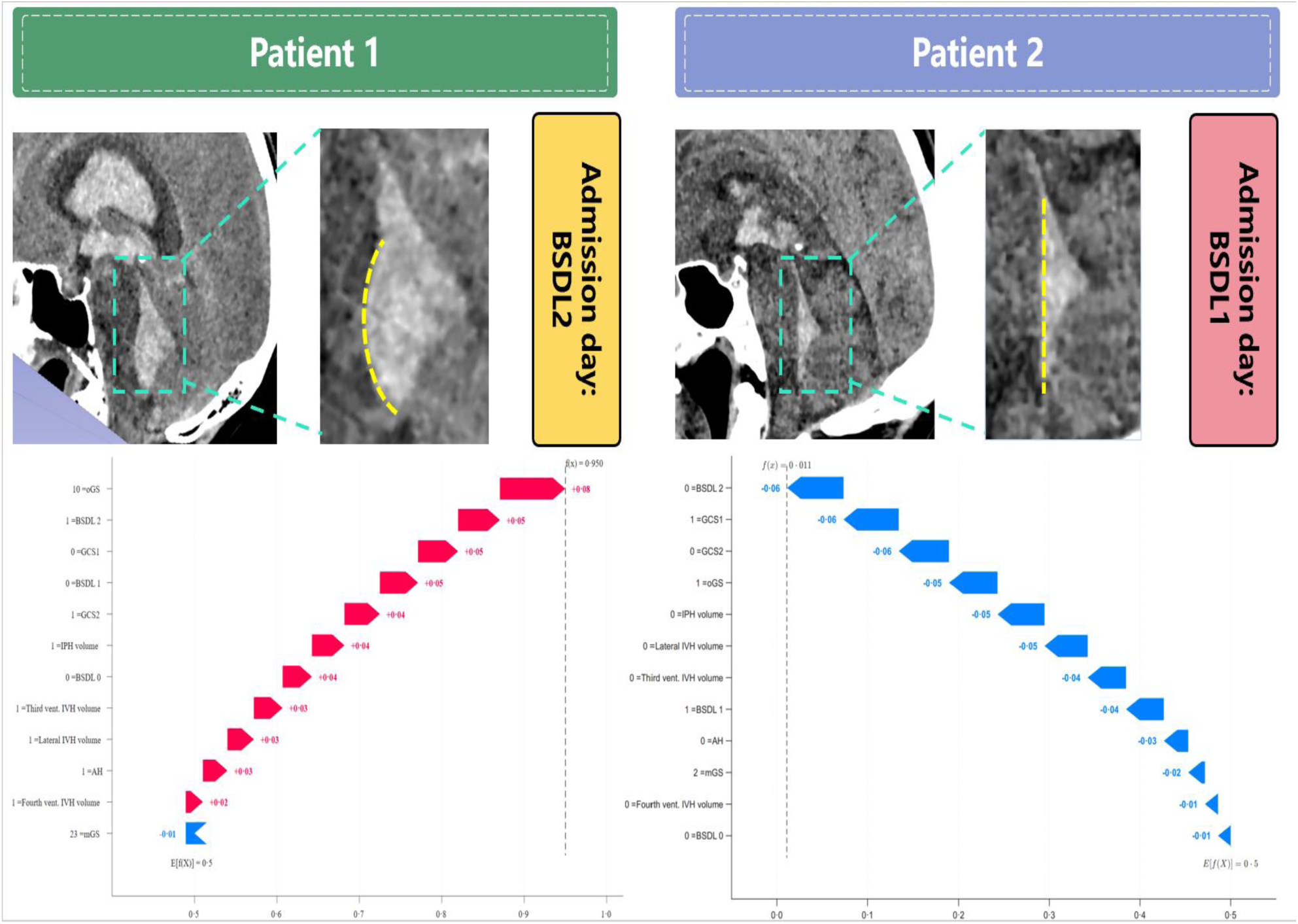
Local-level SHAP waterfall plots for representative cases. Patient 1 is a high-risk case (predicted probability of poor outcome, 0·95); Patient 2 is a low-risk case (predicted probability of poor outcome, 0·011). The upper panels show sagittal admission head CT images and BSDL grade annotations. The lower panels show SHAP waterfall plots. E[f(X)] denotes the model’s baseline predicted probability (0·5), and f(x) represents the final predicted probability. Red arrows indicate positive SHAP contributions, blue arrows indicate negative SHAP contributions, and arrow length reflects contribution magnitude. SHAP=SHapley Additive exPlanations.

In the high-risk case (Patient 1), severe brainstem compression (BSDL grade 2) was evident on admission CT, and the predicted probability of unfavourable outcome was 0·950 (figure 8, left). Key positive SHAP contributors were oGS (+0·080), BSDL grade 2 (+0·050), GCS1 (+0·050), BSDL grade 1 (+0·050), and GCS2 (+0·040), collectively raising the predicted probability well above baseline.

In the low-risk case (Patient 2), mild brainstem compression (BSDL grade 1) was evident on admission CT, and the predicted probability of unfavourable outcome was 0·011 (figure 8, right). Key protective SHAP contributors were absence of BSDL grade 2 (−0·060), GCS1 (−0·060), GCS2 (−0·060), oGS (−0·050), and IPH volume (−0·050), collectively driving the predicted probability well below baseline.

## 4 Discussion

In this multicentre retrospective study, we introduced the BSDL grading system to quantify brainstem compression from fourth-ventricle haematoma. We then developed and externally validated a prognostic model that integrated BSDL grading with TabICLv2 to predict 6-month functional outcomes in patients with IVH. The model achieved superior performance compared with seven other ML algorithms. SHAP analysis identified BSDL grade 2 as an independent predictor of unfavourable outcome, with greater predictive value than conventional imaging features.

Mechanisms of unfavourable outcome after IVH include increased intracranial pressure from haematoma mass effect, acute and chronic hydrocephalus, neurotoxicity from blood degradation products, and direct compression of adjacent brain structures — most importantly, the brainstem ^2,10,19–20,22^. Fourth-ventricle haematoma causes not only mechanical compression but also disruption of the reticular activating system, cardiovascular regulatory centres, and respiratory centres within the brainstem. These effects impair consciousness, destabilise blood pressure, and disrupt respiratory rhythm, directly contributing to unfavourable outcomes.

The importance of fourth-ventricle haematoma in determining IVH outcomes has been recognised for decades. Since Shapiro et al.^29^ first reported fourth-ventricle dilatation as a predictor of poor prognosis in patients with IVH, subsequent studies have confirmed that fourth-ventricle haemorrhage with brainstem compression is a consistent risk factor for unfavourable outcome. Mo et al.^8^ demonstrated that haematoma volume independently predicts unfavourable outcomes, and that posterior fossa craniotomy may improve outcomes in patients with brainstem compression. In aneurysmal subarachnoid haemorrhage, Overstijns et al.^20^ reported an 89% unfavourable outcome rate associated with fourth-ventricle casting and brainstem compression, with aggressive surgical evacuation conferred substantial benefit. However, these studies mainly relied on fourth-ventricle size, haematoma volume, or subjective visual assessment, and lacked a standardised, reproducible quantitative tool, which might explain the controversy in previous findings.

To address this gap, we developed the BSDL grading system, a novel semi-quantitative tool that uses sagittal CT reconstruction and the dorsal brainstem contour as an anatomical reference to assess fourth-ventricle haematoma–induced brainstem compression. BSDL grade 2 was present in 47·1% of patients with unfavourable outcomes versus 4·5% with favourable outcomes; SHAP analysis confirmed it as the primary outcome driver, exceeding the contributions of IPH volume, oGS, mGS, and all individual ventricular haematoma volumes. A clear dose–response relationship between BSDL grade and unfavourable outcome was identified, which may mechanistically explain why prior approaches based solely on haematoma volume have proved insufficient. Within the narrow fourth ventricle, haematomas of equivalent volume can exert variable focal compression on the brainstem depending on spatial distribution. Although oGS and mGS quantify overall ventricular haematoma burden^2,7,19^, they cannot capture this focal effect. This explains why the SHAP contributions of mGS and oGS were lower than that of BSDL grade 2. In clinical practice, BSDL grading could facilitate risk-stratified management of IVH. For patients with BSDL grade 0, conservative management may be preferred, avoiding unnecessary invasive intervention. For those with grade 1, external ventricular drainage (EVD) is recommended, with close monitoring of intracranial pressure and consciousness for haematoma expansion causing brainstem compression. For patients with grade 2, EVD combined with intraventricular fibrinolysis may offer limited benefit, as these interventions fail to rapidly decompress the posterior-fossa mass effect. When surgical criteria are met, early craniotomy or neuroendoscopic evacuation of the fourth-ventricle haematoma should be considered to relieve brainstem compression and improve outcomes. Current treatment paradigms lack criteria for fourth-ventricle haematoma evacuation; BSDL grading addresses this gap by identifying patients with severe brainstem compression who may benefit from early decompression. Prospective randomised controlled trials are needed to validate BSDL grading in guiding surgical decisions and supporting broader clinical implementation.

The GCS, a well-established predictor of outcome after sICH ^9,13,15–17^, ranked second among all features by mean absolute SHAP value, confirming its importance after BSDL grade 2. The two GCS strata showed divergent prognostic effects: low GCS (3–8) strongly predicted unfavourable outcome, whereas high GCS (9–15) was associated with favourable prognosis, a pattern reflecting the model’s capacity to distinguish prognostic effects across levels of consciousness impairment. These findings are in line with Morotti et al.^27^, who reported that the severity of consciousness impairment correlates positively with haematoma expansion and poor outcome in sICH, and underscore the clinical importance of consciousness assessment in prognostic stratification for IVH. Unlike the GCS, which reflects irreversible neurological impairment, BSDL grade 2 represents a mechanically reversible insult—brainstem compression by the fourth-ventricle haematoma—amenable to surgical evacuation.

IPH volume, ventricular haematoma volumes, and AH were also confirmed as independent predictors of unfavourable outcome, consistent with prior studies^5,10,27–28,30^. Lateral IVH volume contributed more to outcome prediction than third-or fourth-ventricle volumes, concordant with García-Pérez et al.^2^, who identified lateral ventricular haematoma as the most important prognostic factor. The underlying mechanism might be that the lateral ventricles, being the largest compartments of the ventricular system, accommodate greater haematoma volumes that more readily elevate intracranial pressure and obstruct cerebrospinal fluid circulation, thereby aggravating secondary brain injury. AH, a common and severe complication of IVH, was independently associated with adverse outcomes, a finding that corroborates our team’s previous work^22^. This suggests that early recognition and prompt treatment of hydrocephalus may improve functional prognosis.

In recent years, ML has been increasingly applied to outcome prediction after intracerebral haemorrhage. However, most studies have relied on conventional algorithms, lacked IVH-specific models, and rarely performed rigorous external validation^13,15–17^. Focusing on IVH, we proposed BSDL grading as a novel imaging indicator and used TabICLv2, an algorithm optimised for tabular data, to develop a predictive model. Multicentre validation demonstrated that TabICLv2 outperformed comparator models and exhibited greater stability in heterogeneous real-world clinical samples, meeting TRIPOD+AI criteria for AI-based clinical prediction model evaluation and generalisability ^24^.

Interpretability is an essential prerequisite for the clinical translation of ML models. We applied SHAP to dissect the decision logic of TabICLv2 at both global and individual levels. SHAP analysis quantified the relative contribution of each predictor, verified the dose–response relationship between BSDL grade and outcome, and elucidated the limitations of relying solely on haematoma volume for prognostic assessment. Individual waterfall plots visually display each patient’s key outcome drivers, helping clinicians to identify core risk factors and providing an objective basis for individualised treatment decisions and clinician–patient communication. This approach effectively addresses the “black-box” problem inherent to ML models.

Despite the promising results, our study has some limitations. First, its retrospective design may have introduced selection bias, despite strict inclusion criteria and consecutive enrolment. Second, enrolment from nine Chinese centres limits geographical and ethnic generalisability; future studies in diverse populations are needed to verify cross-population generalisability. Third, BSDL grading relies on manual delineation, introducing subjectivity; development of automated segmentation and grading algorithms would enhance assessment efficiency and reduce inter-observer variability. Fourth, neuroinflammatory biomarkers and genetic factors were not included, the incorporation of which might enhance predictive accuracy. Fifth, treatment strategies were recorded but not analysed for interaction effects with BSDL grade. Future work should investigate whether BSDL-based risk stratification can guide surgical timing and intervention selection to optimise clinical decision-making.

In conclusion, we introduced BSDL grading and developed the first IVH prognostic model by integrating the TabICLv2 algorithm. BSDL grade 2 emerged as the primary predictor of unfavourable outcome, demonstrating the prognostic importance of mechanical brainstem compression from fourth-ventricle haematoma. This model may guide individualised treatment decisions and rational allocation of healthcare resources. Prospective multicentre studies are needed to validate BSDL-based decision-support systems and assess clinical impact.

## Data Availability

The de-identified participant data, including demographic information, imaging data, and outcome data, that support the findings of this study are available from the corresponding author (Hua Feng) upon reasonable request. Data will be shared with researchers who provide a methodologically sound proposal for the purpose of replicating the results, subject to approval by the institutional review board and execution of a data use agreement. The model development and analysis code are available from the corresponding author upon reasonable request.

## Contributors

CF, SD, YZ, RH and HF conceived and designed the study. CF, SD, LM, SH, JH, YY, XW, YW, YH, and RH were responsible for patient recruitment, clinical data acquisition, and follow-up. SL and YZ developed the analytical algorithms, performed the statistical analysis, and validated the computational models, with methodological input from HF. CF, SD and YZ interpreted the results and drafted the initial manuscript. RH and HF critically reviewed, edited, and approved the manuscript. CF, SD, YZ, SL and RH directly accessed and verified the individual participant-level data reported in the manuscript. All authors had direct access to all data supporting the results of this study. All authors read and approved the final manuscript and take responsibility for the decision to submit for publication.

## Data sharing

De-identified individual participant data, including demographic information, imaging data, and outcome data, will be made available upon reasonable request to the corresponding author (Hua Feng) after publication. Requesters will be required to submit a detailed research protocol, an institutional review board-approved analysis plan, and documentation of data security compliance. Data sharing will proceed only after a data use agreement has been executed and all required approvals have been verified. The study protocol, statistical analysis plan, and informed consent form are also available upon request. The algorithm source code used in this study is available from the corresponding author upon reasonable request. All shared data will be de-identified and provided for non-commercial research purposes only.

## Acknowledgments

We extend our sincere gratitude to the Ethics Committee of the First Affiliated Hospital of the Army Medical University for their ethical oversight and support throughout this study. We are also grateful to the participants and their families for their invaluable contributions to advancing neurological research.

## AI declaration

During the preparation of this work, the author(s) did not use any generative AI or AI-assisted technologies.

